# Biopsy-free prediction of prostate cancer aggressiveness using deep learning and radiology imaging

**DOI:** 10.1101/2019.12.16.19015057

**Authors:** Pegah Khosravi, Maria Lysandrou, Mahmoud Eljalby, Matthew Brendel, Qianzi Li, Ehsan Kazemi, Josue Barnes, Pantelis Zisimopoulos, Alexandros Sigaras, Camir Ricketts, Dimitri Meleshko, Andy Yat, Timothy D. McClure, Brian D. Robinson, Andrea Sboner, Olivier Elemento, Bilal Chughtai, Iman Hajirasouliha

## Abstract

Magnetic Resonance Imaging (MRI) is routinely used to visualize the prostate gland and manage prostate cancer. The Prostate Imaging Reporting And Data System (PI-RADS) is used to evaluate the clinical risk associated with a potential tumor. However the PI-RADS score is subjective and its assessment varies between physicians. As a result, a definite diagnosis of prostate cancer requires a biopsy to obtain tissue for pathologic analysis. A prostate biopsy is an invasive procedure and is associated with complications, including hematospermia, hematuria, and rectal bleeding.

We hypothesized that an Artificial Intelligence (AI) can be trained on prostate cases where both imaging and biopsy are available to distinguish aggressive prostate cancer from non-aggressive lesions using MRI imaging only, that is, without the need for a biopsy.

Our computational method, named AI-biopsy, can distinguish aggressive prostate cancer from non-aggressive disease with an AUC of 0.855 and a 79.02% accuracy. We used Class Activation Maps (CAM) to highlight which regions of MRI images are being used by our algorithm for classification, and found that AI-biopsy generally focuses on the same regions that trained uro-radiolosts focus on, with a few exceptions. In conclusion, AI-biopsy provides a data-driven and reproducible way to assess cancer aggressiveness from MRI images and a personalized strategy to reduce the number of unnecessary biopsies.

## Introduction

Prostate cancer is the most commonly diagnosed cancer in adult men^1^. Distinguishing patients with aggressive (tumor tissue growing faster) and non-aggressive (tumor tissue growing slowly) forms of prostate cancer is important because the former needs more aggressive treatment while the latter may only necessitate monitoring. Indeed early detection and intervention of aggressive prostate cancer improves survival rate^2,3^. In addition, accurate diagnosis prevents over-treatment^4^.

Magnetic Resonance Imaging (MRI) is used by radiologists to visualize abnormalities within the prostate gland. The European Society of Urogenital Radiology (ESUR) established the Prostate Imaging Reporting And Data System (PI-RADS), a standardized guideline for interpretation and reporting prostate MRI for the purpose of harmonizing MRI use^5,6^. PI-RADS is designed to improve detection, localization, characterization, and risk stratification in patients with suspected cancer^7^. PI-RADS uses subjective features such as lesion shape and margins for categorization of prostate cancer^8^. PI-RADS categories range from one to five and higher-grade lesions have higher PI-RADS assessment categories (PI-RADS categories 4 and 5)^9^. Although PI-RADS has been found to be effective in evaluating the clinical risk associated with prostate cancer^10^, it is subjective and relies on visual assessment^11^. As a result, histology assessment based on prostate biopsies remains the standard approach for assessing prostate cancer aggressiveness.

There are currently two main scores used to assess histology slides for prostate cancer aggressiveness. The Gleason Score (GS) is the most commonly used prognostic score to predict the clinical status of prostate cancer (aggressive and non-aggressive prostate cancer) based on biopsy material. GS describes how much the tissue from a biopsy looks like healthy tissue (lower score) or abnormal tissue (higher score)^12-14^. GS is a sum of the primary and secondary patterns with a range of 3 to 5. Thus, tumors are scored as a GS ranging from 6 (3 + 3) to 10 (5 + 5). Another score, Grade Group (GG), subdivides prostate cancer into five categories using pathological characteristics^15,16^. Pathologists use either of these scores in routine clinical practice. Although a biopsy usually provides a definitive diagnosis of prostate cancer, patients undergoing prostate biopsy may experience incorrect staging and complications such as infection and sepsis that it is associated with life-threatening organ dysfunction and death^17^.

In this paper, we hypothesize that prostate cancer aggressiveness can be predicted directly from MRI images using machine learning techniques, perhaps reducing or even removing the need for a tissue biopsy. In recent years, machine learning and especially deep learning approaches have been applied to a variety of medical problems^18-20^, such as lung cancer subtype diagnosis using pathology images^21^, assessing human blastocyst quality after in vitro fertilization^22^, and prostate cancer classification by MRI images^23^. In the latter study^23^, the authors used deep learning and non-deep learning algorithms to classify benign prostate from prostate cancer using axial 2D T2-weighted MRI images of 172 patients^23^. They were able to distinguish benign from cancer lesions with the Area Under Curve (AUC) of 0.84 and 0.70, respectively^23^. In another related study, Kwon *et al*. described a radiomics-based approach to classify clinically significant lesions in multi-parametric MRI (mp-MRI) using three feature-based methods: regression trees, random forests, and a regularization techniques for simultaneous estimation and variable selection (adaptive LASSO). Random forest achieved highest performance with an AUC of 0.82^24^. Recently Rubinstein *et al*. used an unsupervised deep learning method to detect and localize prostate tumors in PET/CT images^25^. This study demonstrated the utility of feature selection using deep learning methods in finding tumors with AUC of 0.899^25^.

Despite these related attempts, currently, there are no robust approaches that can serve as an alternative to prostate biopsy for the detection of prostate cancer aggressiveness. Here, we introduce an AI-based computational technique that uses MRI imaging data as input and recognizes aggressive prostate cancer from non-aggressive forms as defined by pathology assessment such as Gleason Score and Grade group. While the training uses pathology assessment obtained from a biopsy, our objective is to train a predictive model that can eventually assess MRI images without the need of a biopsy.

To train and test our predictive models, we utilize three publicly available datasets as well as data generated in our radiology practice at Weill Cornell Medicine. Using supervised and unsupervised analyses, we curated a dataset which contains thousands of high-quality MRI prostate images. These images originate from four different databases with pathology labels, and form the largest collection of de-identified human MRI prostate images ever created so far. We used this large-scale imaging dataset to train and assess several Deep Neural Network (DNN) models to classify aggressive and non-aggressive prostate cancer. These models automate prostate cancer diagnosis directly from MRI images, thus empowering uro-radiologists to provide earlier and reduced-risk diagnosis.

## Results

In total, we used 26,257 MRI images from 376 patients, obtained from public resources (The Cancer Imaging Archive (TCIA)) and our medical center (Department of Urology at Weill Cornell Medicine). All 376 patients were labeled with Gleason score or Grade group. Some datasets (e.g., PROSTATE-DIAGNOSIS) used Gleason scores, others (e.g., PROSTATEx) use Grade group. Gleason scores and Grade groups were mapped onto one another as shown in Table 1 based on the National Comprehensive Cancer Network (NCCN) guidelines (see Methods).

**Table 1:** Grade group and Gleason score and their association with aggressiveness of prostate cancer. These two different systems are mapped together based on the below table for this project.

| Grade group | Gleason score | Combined Gleason score | Aggressiveness degree |
| --- | --- | --- | --- |
| Grade group 1 | 3 + 3 | 6 | Low risk |
| Grade group 2 | 3 + 4 | 7 | Intermediate risk but closer to low risk |
| Grade group 3 | 4 + 3 | 7 | Intermediate risk but closer to high risk |
| Grade group 4 | 4 + 4, 3 + 5, 5 + 3 | 8 | High risk |
| Grade group 5 | 4 + 5, 5 + 4, 5 + 5 | 9 and 10 | High risk |

Instead of predicting Gleason score or Grade group which contain too many categories for effective machine learning, we grouped patients into four non-overlapping major groups based on Gleason score (either from original dataset or mapped from Grade group). The first two groups, aggressive prostate cancer (n = 49 patients with Gleason score ≥ 8) and non-aggressive prostate cancer (n = 86 patients with Gleason score = 6) with total 135 patients are used for training and validation. Two other groups, intermediate prostate cancer (n = 167 patients with Gleason score = 7), and benign prostate tissue (n = 74 patients) are used as independent test sets (Table 2).

**Table 2:** Characteristics of all four cohorts and the comprised images.

| database | Refer name | Annotated patients | Annotation method | Patients with high risk label (GS ≥ 8) (G4 & G5) | Patients with low risk label (GS = 6) (G1) | Patients with intermediate risk label (GS = 7) (G2 & G3) | Patients with benign label |
| --- | --- | --- | --- | --- | --- | --- | --- |
| Weill Cornell Medicine | WCM | 203 | Gleason Score | 15 | 50 | 64 | 74 |
| PROSTATEx | Ex | 99 | Grade group | 13 | 29 | 57 | 0 |
| PROSTATE-DIAGNOSIS | Dx | 48 | Pathology result | 10 | 7 | 31 | 0 |
| PROSTATE-MRI | MIP | 26 | Pathology result | 11 | 0 | 15 | 0 |
| Total | Public and WCM | 376 | Grade group | 49 | 86 | 167 | 74 |

MRI is able to image in various standard orthogonal imaging planes such as axial images as well as sagittal and coronal images, or any degree in between^26^. Therefore, our datasets contain multiple images from each patient (Figure 1a). Each image is 320×320 pixel resolution in black and white (Figure 1a shows a subset of images for one randomly selected patient). However, not all these layers and rotation of images shows the prostate gland. Therefore, we use a preprocessing method to identify informative images, that is, images whose orientation or focal depth are most helpful for determining aggressive vs. non-aggressive classification (Figure 1).

**Figure 1:**
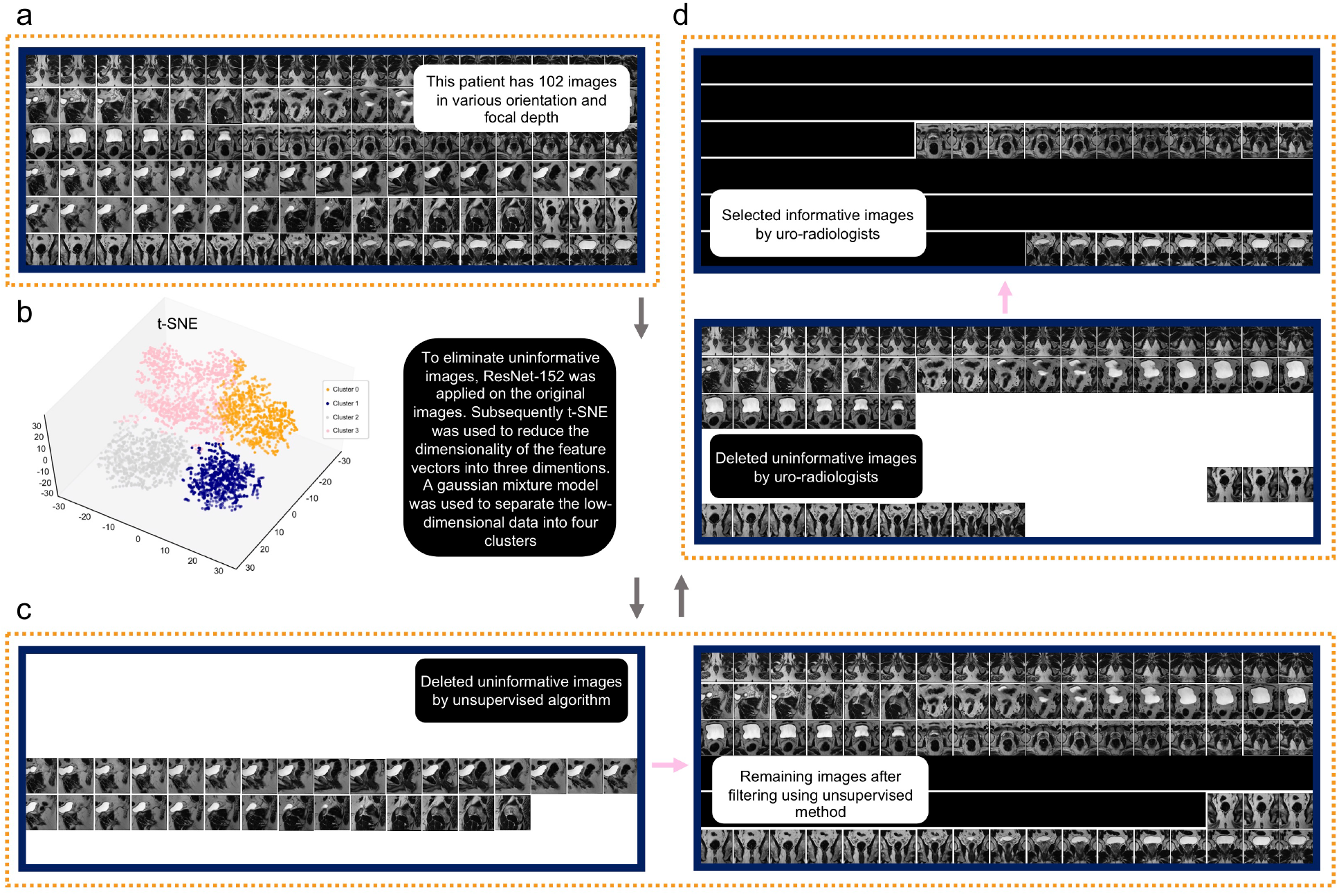
Preprocessing approach to generate informative images for each patient. (a) Each patient’s MRI series contains different orientation and focal depth of images; here for example the patient has 102 images; (b) After feature extraction using a Convolutional Neural Network (CNN), the t-SNE algorithm was used to reduce feature dimension. A gaussian mixture model was used to separate the low-dimensional data into four clusters; (c) An unsupervised algorithm detects some of the most uninformative images (e.g. these 31 images) whose orientation or focal depth are not useful for aggressive vs. non-aggressive classification; (d) Subsequently, an expert uro-radiologist trimmed remaining images (= 71 images) of each patient manually and selected informative images (e.g., these 19 images) with useful orientations and focal depths.

### Unsupervised method extracts uninformative images

We initially reasoned that unsupervised analysis of MRI images followed by examination of the main image cluster may help identify many of the informative images or eliminate uninformative ones. We used a state-of-the-art deep learning algorithm, ResNet-152^27^, pretrained on ImageNet to extract features from the original images (10,904 images for 135 patients with GS = 6 and GS ≥ 8) as described elsewhere^25^. This resulted in a feature vectors containing 2,048 floating point values per image. t-Distributed Stochastic Neighbor Embedding (t-SNE)^28^, a non-linear algorithm for dimensionality reduction, was then used to reduce the dimensionality of the 2,048-dimensional vectors into three dimensions^29^ (Figure 1b).

From there a Gaussian Mixture Model (GMM) was applied to the data, and four clusters (components) were set as the input. We consider four clusters because the configuration with four clusters is the most conservative based on the amount of information included (=biggest possible number of clusters) and on the stability of the fitting procedure (data not shown).

Then, we asked expert uro-radiologists to examine representative images for each cluster. Although the unsupervised clustering method could not classify images into clinically relevant groups (i.e. all clusters contains aggressive and non-aggressive images), images that were deemed informative by a uro-radiologist clustered into three distinct clusters. The fourth cluster largely contained uninformative images (Figure 1c). Therefore, we eliminated the single cluster of images (cluster 1 in Figure 1b) that contains all uninformative images (n = 3,170). To discover additional uninformative images, we asked our uro-radiologists who were blinded to PI-RADS score and Gleason score to manually select images (out of the remaining 7,734 images) whose orientation or focal depth they considered most useful for determining aggressive or non-aggressive status (Figure 1d). 1,619 images were selected using this approach.

### Deep Neural Network classify MRI images

The aggressive/non-aggressive images (Gleason score ≥ 8 and Gleason score = 6, respectively) for 135 patients include a total of 10,904 images (original dataset). Out of these, 7,734 were automatically selected based on clustering described in the previous section (clustered dataset) and 1,619 were manually selected (manually-selected dataset) (Table 2).

Based on these initial datasets, we trained Inception-V1 models using the two prostate status groups at both ends of the spectrum, i.e., Gleason score = 6 (non-aggressive) and Gleason score ≥ 8 (aggressive). We used a transfer learning strategy to train the Inception-V1 architecture^30^, where we initially performed fine-tuning of the parameters for all of the layers. We used 5,000 steps for training the DNNs and subsequently evaluated the performance of our DNNs using a randomly selected independent test set with 1323, 958, and 224 images from the original (Model 1), clustered (Model 2), and manually-selected (Model 3) datasets. Table 3 summarizes the different groups, models and dataset sizes.

**Table 3:** Characteristics of all trained models and the comprised images.

| **Models** | **Data resources** | **Total number of images in test set** | **Number of images in aggressive class (train set)** | **Number of images in non-aggressive class (train set)** |
| --- | --- | --- | --- | --- |
| Model1 (mix-original) | WCM and public | 1281 images (18 patients) | 2742 images (40 patients) | 6881 images (77 patients) |
| Model2 (mix-cluster) | WCM and public | 922 images (18 patients) | 2137 images (40 patients) | 4675 images (77 patients) |
| Model3 (mix-manual) | WCM and public | 224 images (18 patients) | 582 images (40 patients) | 813 images (77 patients) |
| Model4 (WCM-manual) | WCM | 73 images (6 patients) | 159 images (13 patients) | 508 images (46 patients) |
| Model5 (public-manual) | Public | 151 images (12 patients) | 423 images (27 patients) | 305 images (31 patients) |
| Model6 (AI-biopsy) | WCM and public | 224 images (18 patients) | 728 images (51 patients) | 813 images (77 patients) |

Our results showed that the trained algorithm was able to distinguish aggressive from non-aggressive images with AUCs of 0.754, 0.760, and 0.853 for Model 1, Model 2, and Model 3 images, respectively (Figure 2a). The performance of Model 2 (accuracy = 71.26%, sensitivity = 76.65%, specificity = 67.74%) in detection of aggressive cases (aggressive predictive value = 60.78%) (Figure 2b) is slightly higher than Model 1 (aggressive predictive value = 54.99%), hinting that detecting and removing uninformative images improves performance (accuracy = 71.04%, sensitivity = 75.58%, specificity = 68.74%). Model 3 has the highest accuracy for classification of aggressive cases (aggressive predictive value = 71.43%), suggesting that manual selection of informative images outperforms (accuracy = 75.45%, sensitivity = 77.67%, specificity = 73.55%) automated selection. When separating images from public and WCM sources in the test set, we find that Model 3 can classify aggressive and non-aggressive images by AUC of 0.821 (public) and 0.788 (WCM) (Figure 2c).

**Figure 2:**
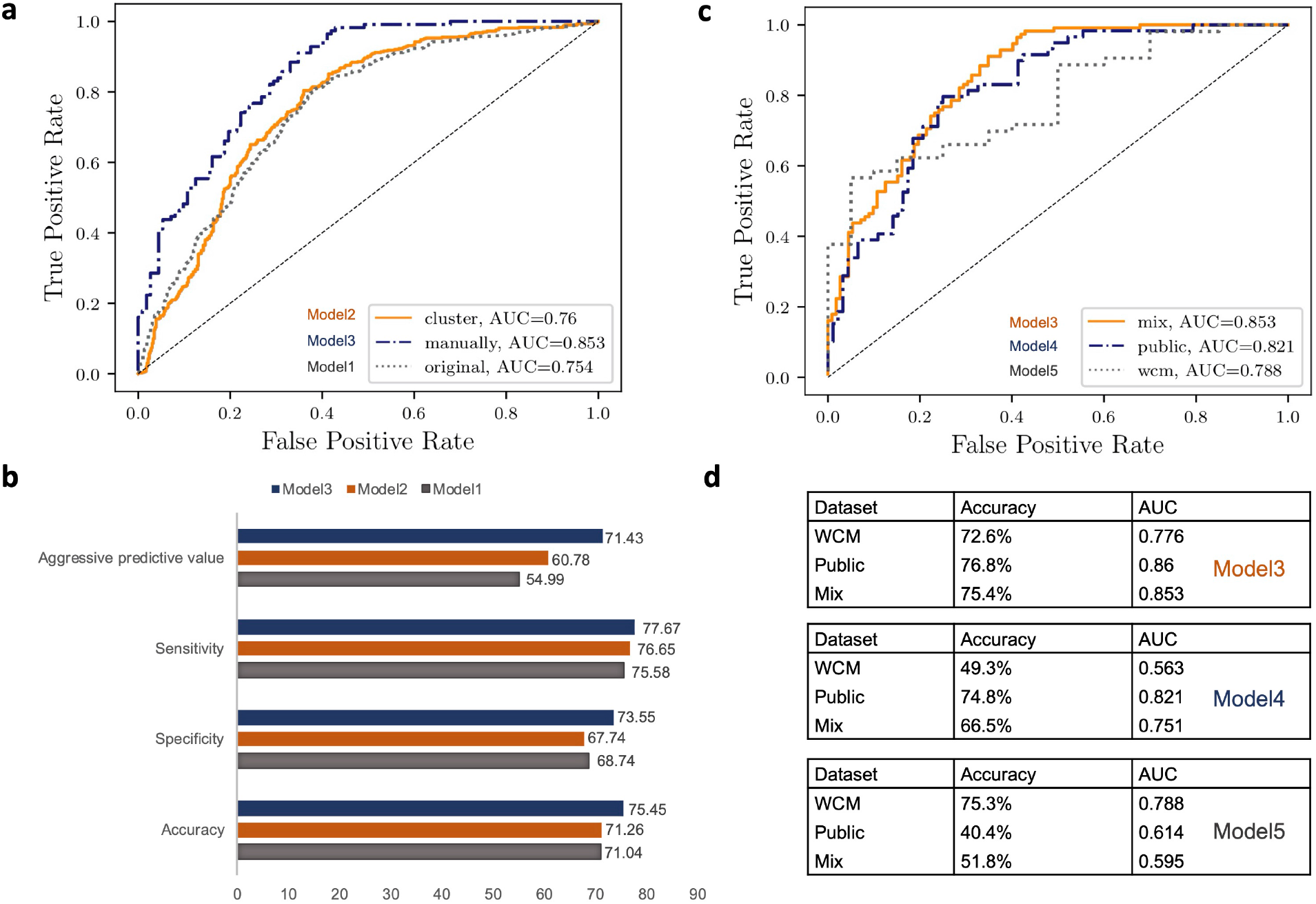
Performance of various trained models using different test sets. (a) Comparing Model 1, Model 2, and Model 3 performance using test data comes from combined (public and WCM) shows that manually selection of images are the best method to detect the informative images; (b) While training the algorithm by the original images is not accurate for detecting aggressive cases, the manually selection method is the best one for this purpose; (c) Integration of data from various datasets increase the performance of algorithm. The figure shows Model 3, Model 4, and Model 5 that are trained and tested using combined WCM and Public data, WCM only data, and public only data, respectively; (d) Model 3 versus Model 4 and Model 5 for classification of prostate states through various databases. The figures show the Model 3, Model 4, and Model 5 that are trained using Mix data, WCM only data, and public only data, respectively. The models also tested by Mix data, WCM only data, and public only data, respectively.

We sought to determine whether training predictive models from a single source would improve or decrease performance. We thus trained Inception-V1 models using images from the manually-selected dataset, focusing on WCM data only (Model 4), and public data only (Model 5; Figure 2d).

We then tested the trained models with blind test sets. We observed low accuracy (Figure 2d) in both Model 4 (= 66.5%) and Model 5 (= 51.8%). This is mainly due to a strong imbalance of aggressive vs. non-aggressive cases: there was a higher proportion of images of aggressive prostate cancer images in public databases while the WCM database contained mainly non-aggressive prostate cancer images (Table 2). Therefore, integration of images from various databases (Model 3) make the classes more balanced, thus increasing the accuracy (= 75.4% accuracy) of the trained Model (Figure 2d).

### Deep learning algorithm outperforms PI-RADS for classification

To further evaluate Model 3, we also tested it using images from 74 patients with benign prostate tissue (not a malignant tumor) obtained from the WCM database and not included in the original training set. We expected the trained algorithm to classifies all the patients as non-aggressive patients. Indeed the result showed that the Model 3 classify the benign status with an 86.5% accuracy in assessing images.

We also assessed how Model 3 would assess intermediate class images (Gleason score = 7). Here we take advantage of the granularity of grade groups in that intermediate class, divided into Grade group = 2 (less aggressive) and 3 (more aggressive).

Model 3 correctly recovered 79.7% of these labels (Figure 3a). We wondered if PI-RADS scores had comparable accuracy. A single PI-RADS score is available for each patient in our database. To measure the accuracy of Model 3 for individual patients (as opposed to images), we used a simple voting system across multiple images for each patient. If the majority of the images from the same patients were predicted to be aggressive, then the final label of the patient was considered aggressive. Of the intermediate class, PI-RADS score distinguished 52 patients as more aggressive (PI-RADS score = 4 and 5) and 10 patients as less aggressive (score = 1, 2, and 3). When compared to pathology, PI-RADS score predict 37.5% of the patients correctly in this intermediate group.

**Figure 3:**
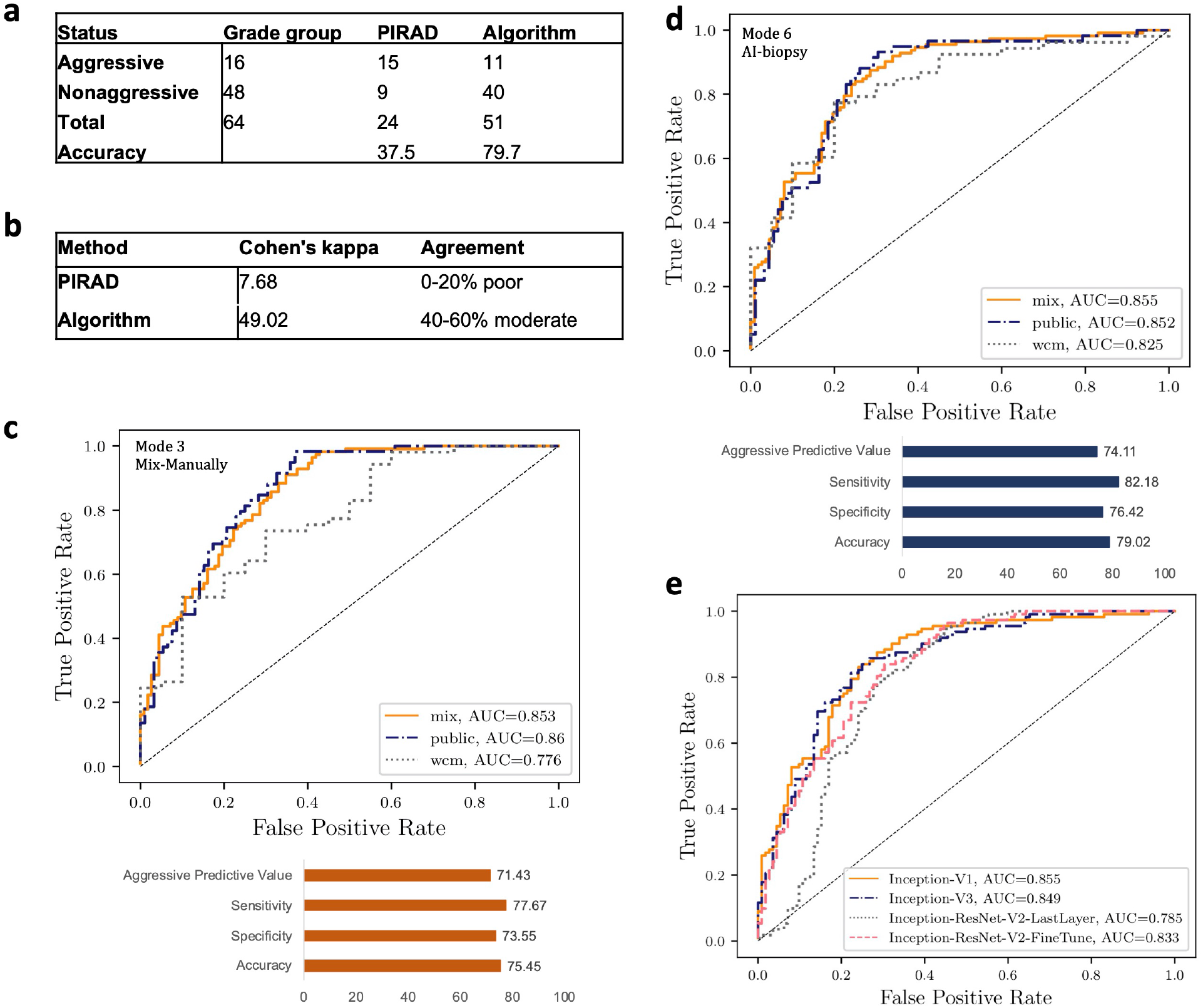
Comparing the result of PI-RADS and the trained models. (a) The algorithm (Model 3) has a higher accuracy in predicting the intermediate class compared to the performance of PI-RADS score for the same diagnosis; (b) The algorithm result is in more agreement with the pathology result compared to PI-RADS; (c) The performance of the Model 3 in detecting aggressive and non-aggressive cases; (d) Model 6 (AI-biopsy) is more accurate in detecting aggressive prostate cancer status compared to Model 3; (e) Using more complex algorithms such as Inception-V3 and Inception-ResNet-V2 cannot increase the performance of the trained algorithm using Inception-V1.

To assess the agreement between Model 3 and PI-RADS scores, we also calculated the Cohen’s kappa for PI-RADS and Model 3 in comparison to the pathology results in this intermediate group (Gleason score = 7, Grade group 2 or Grade group 3). The results showed that while the Model 3 has moderate agreement (= 49.02%) with pathology result, the PI-RADS score estimated by uro-radiologist has poor agreement with pathologists (= 7.68%) (Figure 3b). This suggests that the trained algorithm (Model 3) finds sufficient structure within patients with Gleason Score = 7 to make clinically relevant predictions.

### AI-biopsy is a reliable alternative for biopsy

In previous studies^21^, we showed that increasing the number of images in each class reduces the discordance of machine learning model predictions and pathology results. We started with 582 images for aggressive class and 813 images for the non-aggressive class. To produce a more balanced dataset, we added all those images with intermediate grades that are classified as aggressive cancer using pathology result (Gleason score = 4+3 or Grade group = 3), PI-RADS score (≥ 4), and the Model 3) to the training dataset. We obtained 728 images for aggressive class and 813 images for the non-aggressive class. Then, we again trained Inception-V1 using 5,000 steps on this new dataset (Model 6). Model 6, also called AI-biopsy moving forward (Figure 3d) can classify aggressive vs. non-aggressive classes with a higher performance (accuracy = 79.02%, sensitivity =82.18%, specificity = 76.42%) compared to Model 3 (accuracy = 75.45%, sensitivity =77.67%, specificity = 73.55%) (Figure 3c).

We also compared various CNN architectures to assess their performance on classification of prostate MRI images. These included Google’s Inceptions versions 1 and 3, fine-tuning the parameters for all layers of our network, and the ensemble of two the state-of-the-art algorithms (i.e., Inception and ResNet) via fine-tuning all the layers (Inception-ResNet-V2-FineTune) and training the last layer (Inception-ResNet-V2-LastLayer).

As Figure 3e demonstrates, the inception-based architecture networks (V1 and V3) as well as Inception-ResNet-V2 that are fine-tuned for all layers, are consistently superior compared to the Inception-ResNet-V2 trained for last layer (Figure 3e). We also found that using Inception-V3 and Inception-ResNet-V2 did not increase the performance in classifying aggressive and non-aggressive patients compared to Inception-V1.

### Investigation of discriminative localization using deep features analysis

We reviewed the AI-biopsy (Model 6) result for the test set (n = 18 patients). We also analyzed 9 randomly selected intermediate cases not part of the training set, resulting in 27 patients from each of which 1 image was selected at random (n = 27 images). The results indicated the AI-biopsy was correct in predicting 21 out of 27 images from different patients (Figure 4a). We wondered if the disagreement between AI-biopsy and pathologists for the six patients is due to the incorrect pathology labeling or it shows the AI-biopsy did not look at the correct region of the images. To this end, we have to identify which regions of an image are exactly being used by CNNs for discrimination. We used Class Activation Map (CAM)^31^ as introduced by Zhou *et al*.^31^. We used the same procedure as in Zhou *et al*. for generating CAM using Global Average Pooling (GAP) in CNNs. Before the final output layer (softmax) of the AI-biopsy, we performed global average pooling on the convolutional feature maps and used those as features for a fully-connected layer. Given this connectivity structure, we could identify the importance of the image regions by projecting back the weights of the output layer onto the convolutional feature maps (Figure 4b). In parallel, we independently asked an expert uro-radiologist to highlight the region of each image that is important for categorization as aggressive or non-aggressive (Figure 4c). We compared the CAM result (Figure 4b) with uro-radiologist result (Figure 4c) to assess the power of AI-biopsy in detecting the prostate gland region and the reason of disagreement between AI-biopsy and pathology labels in the six cases.

**Figure 4:**
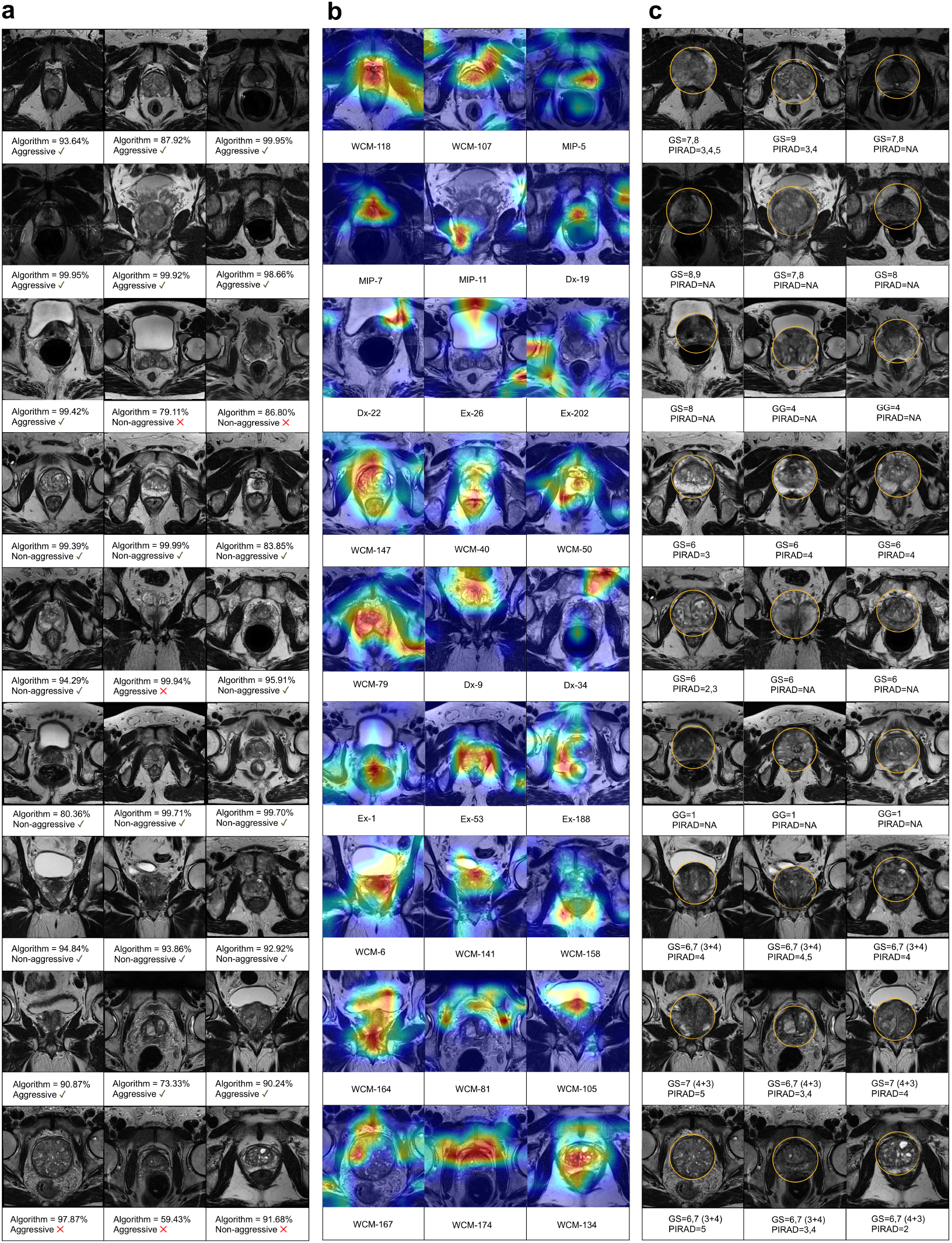
The highlighted prostate gland using CAM and uro-radiologists. (a) The algorithm classifies each images as aggressive or non-aggressive; (b) Deep feature analysis highlights the discriminative regions of the images for various images; (c) An uro-radiologist reviewed some of images and highlighted the important part of each image to show regions of images that are used for discrimination by uro-radiologists.

Applying CAM on AI-biopsy showed that the probability score of AI-biopsy is lower than 90% for three out of the six discordant cases; this indicates that AI-biopsy was relatively uncertain regarding the prediction (Figure 4a). For two out of these three cases, the CAM analysis indicates that the algorithm is not looking at the prostate gland (Figure 4b). In addition, for the remaining three incorrect predicted cases, the AI-biopsy algorithm predicts them as aggressive, aggressive, and non-aggressive, respectively with more than 90% confidence (Figure 4a). The PI-RADS (available for two out of three cases) also confirms the AI-biopsy results in detecting aggressive and non-aggressive cases. The agreement between AI-biopsy and PI-RADS may suggest partially incorrect pathology assessments for these cases.

## Discussion

The early and precise diagnosis of prostate cancer is critical for proper management of patients. Currently a prostate biopsy is the most common method of diagnosis but it carries risks including infection. Prostate MRI is used to improve the biopsy process by identifying the most suspicious area to biopsy. The most commonly used prostate MRI annotation system is the PI-RADS score. PI-RADS is subjective and currently no robust non-invasive method exist leveraging MRI images for determining prostate cancer aggressiveness. To overcome this problem, we sought to determine whether a deep learning method using MRI that are labeled based on histology results could be used instead of biopsy. We proposed a novel CNN-based method, AI-biopsy, to fully utilize MRI images and jointly detect prostate cancer using Gleason score. We trained and validated AI-biopsy using MRI images of 376 patients that are labeled with histopathology information. AI-biopsy achieved an AUC of 0.855 (accuracy = 79.02%, sensitivity =82.18%, specificity = 76.42%) for classification of aggressive vs. non-aggressive prostate cancer (GS ≥ 8 vs. GS = 6). Our method also achieved an accuracy of 79.7% to classify GS ≥ 4+3 vs. GS ≤ 3+4 which was higher than experienced radiologists using PI-RADS (= 37.5%).

Several groups before us have attempted to use different deep learning-based approaches for assessment of prostate cancer aggressiveness with varying degrees of success^32-34^. Cao and colleagues proposed multi-class CNN (named FocalNet) to detect prostate lesion^34^. They used the MRI images of 417 patients to predict Gleason score using an in-house cohort and showed that their method outperformed U-Net^35^ and Deeplab^36^, both of which are CNN-based methods^34^. They trained their model to predict four GS groups such as GS ≥ 7 vs. GS < 7, GS ≥ 4+3 vs. GS ≤ 3+4, GS ≥ 8 vs. GS < 8, and GS ≥ 9 vs. GS < 9. Their result showed that FocalNet achieved AUC of 0.81, 0.79, 0.67, and 0.57, respectively^34^. A recent study^32^ has used Apparent Diffusion Coefficient (ADC), a metric that is correlated with Gleason score and is an essential component of mp-MRI for determining aggressiveness of prostate cancer. They used MRI images of 165 patients and predict high-grade (GS ≥ 7) from low-grade (GS = 6) prostate cancer with AUC of 0.79^32^. In addition, Yuan *et al*. presented a deep learning-based method to classify 123 patients with high-grade cancer (GS = 4 + 3, and 8) and 98 patients with low-grade cancer (GS = 3 + 4, and 6)^33^ based on cropped mp-MRI images. The best performance was obtained using a patch size of 28×28 pixels, which led to classifying the two groups with an AUC of 0.896^33^.

Although these methods achieved good accuracy in assessing prostate cancer aggressiveness, they require several preprocessing steps. Also, they often do not leverage diverse enough datasets. Moreover their data does not cover all Gleason scores. The advantage of our method is that instead of only focusing on predetermined, segmented features to analyze, the entire image of the prostate is assessed, allowing for quantification of all the available data. However, our dataset also has limitations. Our MRI images are labeled using pathology labels that may include inaccurate histologic finding. Moreover the MRI images do not provide the information of the exact shape, location, and size of the lesions. Further study is needed to consolidate the connection between MRI and prostate cancer aggressiveness, particularly with available molecular subtypes of prostate cancer.

In spite of these limitations, our study and training dataset are larger than those used previous investigations that have sought to determine aggressiveness of the prostate cancer. Also, our method covers all Gleason scores ranges (GS = 3+3, 3+4, 4+3, 4+4, 3+5, 5+3, 4+5, 5+4, and 5+5) in addition to benign lesion. Our novel method is developed using thousands of images from various resources in addition to our own WCM database.

Finally, AI-biopsy is automated and does not require any manual augmentations or pre-processing for testing any new images. AI-biopsy can be implemented in clinical practice by providing uro-radiologists a straightforward platform to use without requiring sophisticated computational knowledge.

## Methods

### Providing databases from four different resources

This study included 203 patients from our own Urology center (2015/02–2019/03). We referred to this dataset as WCM throughout this manuscript. This study used fully de-identified data and was approved by the Institutional Review Board at Weill Cornell Medicine (IRB number: 1601016896). The images were provided using 3-T mp-MRI with various orientation and layers, representing about 15,000 images (320×320 pixels). We also used three external public datasets obtained from TCIA including the PROSTATEx Challenge with 99 patients, PROSTATE-MRI which comprises a total of 26 patients of prostate MRI, and PROSTATE-DIAGNOSIS of Prostate cancer MRIs (48 patients).

The patients in each database are labeled for Gleason score (6 to 10) or Grade group (1 to 5) by pathologists and PI-RADS (1 to 5) by uro-radiologists (if it is available).

The Gleason Score is the grading system used to determine the aggressiveness of prostate cancer^37^. This grading system can be used to choose appropriate treatment options. The Gleason scores ranges from 6-10 and describes how much the cancer from a biopsy looks like healthy tissue (lower score) or abnormal tissue (higher score). Since prostate tumors are often made up of cancerous cells that have different grades, two grades are assigned for each patient. A primary grade is given to describe the cells that make up the largest area of the tumor and a secondary grade is given to describe the cells of the next largest area. For instance, if the Gleason Score is written as 3+4=7, it means most of the tumor is grade 3 and the next largest section of the tumor is grade 4, together they make up the total Gleason Score. If the cancer is entirely made up of cells with the same score, the grade for that area is counted twice to calculate the total Gleason Score. The higher the Gleason Score, the more likely the cancer will grow and spread quickly. Score of 6 describes cancer cells are likely to grow slowly. Scores of 8 or higher describe high risk for aggressive cancer that are likely to spread more rapidly. These cancers are often referred to as poorly differentiated or high grade. Finally, score 7 indicates intermediate risk.

Although Gleason Score is routinely used to determine the prostate cancer risk, some resources report the aggressiveness of prostate cancer with a grading system which shows the prostate cancer aggressiveness based on grade group. Grade group uses 1 to 5 different grades (Table 1). In Table 1 we map the cancer Grades groups with Gleason scores for those images that shows the aggressiveness with different system. This table was provided and simplified based on the National Comprehensive Cancer Network (NCCN) guidelines version 4.2018 prostate cancer^38^. In addition, we labeled those images without label based on their pathology report (as excel file or histological whole slide image).

Characteristics of all four databases and their images are summarized in Table 2.

This study presents a framework to classify MRI images based on Gleason scores and Grade group and map those scores and grades to aggressive and non-aggressive prostate cancer.

Finally, we divided the images into training, validation, and test groups. We allocated 70% of the images to the training group and the remaining 30% to the validation and test groups. The training, validation, and test sets did not overlap. Characteristics of training and test sets of trained models and the comprised images are summarized in Table 3.

### Algorithm Architectures and Training Methods

For the unsupervised learning process prior to model training, feature extraction was performed using Resnet-152^27^ pretrained on ImageNet in PyTorch. The last fully-connected layer was removed, such that the output of a single image was 2048 ⨯ 1. From there t-SNE^28^ was used for dimensionality reduction to three dimensions, and a gaussian mixture model using the scikit-learn package in python was used to cluster image features into four clusters based on manual inspection.

For supervised learning, we used Google’s Inception-V1^30^ and Inception-V3^39^ architectures, which offers an effective run-time and computational cost. To train this architecture, we used transfer learning and pretrained network on the ImageNet dataset. We then fine-tuned all outer layers using the MRI images obtained from WCM and public resources. We also used the ensemble architecture, Inception-Resnet-V2 for supervised learning^40^. We trained the latter architecture using two strategies: fine-tuned all outer layers and last layer training. To implement our framework called AI-biopsy, we used Tensorflow version 1.4.028 and the TF-Slim Python library for defining, training, and evaluating models in TensorFlow. Training of our DNN method was performed on a server running the SMP Linux operating system. This server is powered by four NVIDIA GeForce GTX 1080 GPUS with 8 GB of memory for each GPU and 12 1.7-GHz Intel Xeon CPUs^41^.

### Evaluation of the Developed Method

To evaluate the performance of our method, we used an accuracy measure, which is the fraction of correctly identified images. To assess the performance of different algorithms, Receiver Operating Characteristics (ROCs) were estimated. The ROC curve is depicted by plotting the True Positive Rate (TPR) versus the False Positive Rate (FPR) at various threshold settings. The accuracy is measured by the area under the ROC curve (AUC)^42,43^. We also address other measures such as Cohen’s kappa^44^ which is a popular way of measuring the accuracy of presence and absence predictions. The kappa statistic ranges from -1 to +1, where +1 indicates perfect agreement and values of zero or less indicate a performance no better than random^44^.

## Data Availability

The links to the publicly available data sets from TCIA are available below:
The public data obtained from TCIA: https://www.cancerimagingarchive.net/
PROSTATEx
https://wiki.cancerimagingarchive.net/display/Public/SPIE-AAPM-NCI+PROSTATEx+Challenges
PROSTATE DIAGNOSIS
https://wiki.cancerimagingarchive.net/display/Public/PROSTATE-DIAGNOSIS
PROSTATE MRI
https://wiki.cancerimagingarchive.net/display/Public/PROSTATE-MRI

https://wiki.cancerimagingarchive.net/display/Public/SPIE-AAPM-NCI+PROSTATEx+Challenges

https://wiki.cancerimagingarchive.net/display/Public/PROSTATE-DIAGNOSIS

https://wiki.cancerimagingarchive.net/display/Public/PROSTATE-MRI

## Code availability

The source code and the guideline are publicly available at https://github.com/ih-lab/ai-biopsy. In addition, AI-biopsy is available through a web-based user interface at https://ai-biopsy.eipm-research.org (password protected).

## Acknowledgements

This work was supported by start-up funds (Weill Cornell Medicine) to IH. This work used the Extreme Science and Engineering Discovery Environment (XSEDE) GPU servers through allocation ASC180052 to IH. The authors thank Richard Kneppe, Tom Maiden, John Ruffing, and Hanif Khalak for their assistance with porting and optimization which was made possible through the XSEDE Extended Collaborative Support Service (ECSS) program. PK also thank Sinan Ramazanoglu, and Hamid Mohamadi for their help with code understanding and debugging. IH also acknowledges NVIDIA for the donation of a Titan Xp to support this research through a GPU gift grant.

## Author Contributions

PK, AS, OE, BC, and IH conceived the study. PK, MB, QL, EK, and JB conceived the method and designed the algorithmic techniques. PK, ML, and ME generated the datasets and prepared and labeled the images for various Grades groups and Gleason scores. PK, MB, QL, EK, CR, and DM wrote the codes. PK, ML, MB, and QL performed computational analysis with input from AS, OE, BC, and IH. PZ and AS developed the web interface. TDM and AY reviewed the MRI images and PI-RADS scores. BDR reviewed pathological images, Gleason scores, and Grade groups. PK wrote the paper, and all authors read, edited, and approved the final manuscript.

## References

1 Pilleron, S. et al. Global cancer incidence in older adults, 2012 and 2035: A population-based study. Int J Cancer 144, 49–58, doi:10.1002/ijc.31664 (2019).

2 Hricak, H., Choyke, P. L., Eberhardt, S. C., Leibel, S. A. & Scardino, P. T. Imaging prostate cancer: a multidisciplinary perspective. Radiology 243, 28–53, doi:10.1148/radiol.2431030580 (2007).

3 Lim, H. K., Kim, J. K., Kim, K. A. & Cho, K. S. Prostate cancer: apparent diffusion coefficient map with T2-weighted images for detection--a multireader study. Radiology 250, 145–151, doi:10.1148/radiol.2501080207 (2009).

4 Barrett, T. & Haider, M. A. The Emerging Role of MRI in Prostate Cancer Active Surveillance and Ongoing Challenges. AJR Am J Roentgenol 208, 131–139, doi:10.2214/AJR.16.16355 (2017).

5 Hamdy, F. C. et al. 10-Year Outcomes after Monitoring, Surgery, or Radiotherapy for Localized Prostate Cancer. N Engl J Med 375, 1415–1424, doi:10.1056/NEJMoa1606220 (2016).

6 Bittner, N., Merrick, G. S., Butler, W. M., Bennett, A. & Galbreath, R. W. Incidence and pathological features of prostate cancer detected on transperineal template guided mapping biopsy after negative transrectal ultrasound guided biopsy. J Urol 190, 509–514, doi:10.1016/j.juro.2013.02.021 (2013).

7 Padhani, A. R. et al. Prostate Imaging-Reporting and Data System Steering Committee: PI-RADS v2 Status Update and Future Directions. Eur Urol 75, 385–396, doi:10.1016/j.eururo.2018.05.035 (2019).

8 Krishna, S., Schieda, N., McInnes, M. D., Flood, T. A. & Thornhill, R. E. Diagnosis of transition zone prostate cancer using T2-weighted (T2W) MRI: comparison of subjective features and quantitative shape analysis. European Radiology 29, 1133–1143, doi:10.1007/s00330-018-5664-z (2019).

9 Vargas, H. A. et al. Updated prostate imaging reporting and data system (PIRADS v2) recommendations for the detection of clinically significant prostate cancer using multiparametric MRI: critical evaluation using whole-mount pathology as standard of reference. Eur Radiol 26, 1606–1612, doi:10.1007/s00330-015-4015-6 (2016).

10 Portalez, D. et al. Validation of the European Society of Urogenital Radiology Scoring System for Prostate Cancer Diagnosis on Multiparametric Magnetic Resonance Imaging in a Cohort of Repeat Biopsy Patients. Eur Urol 62, 986–996, doi:10.1016/j.eururo.2012.06.044 (2012).

11 Khalvati, F., Zhang, Y., Le, P. H. U., Gujrathi, I. & Haider, M. A. PI-RADS Guided Discovery Radiomics for Characterization of Prostate Lesions with Diffusion-Weighted MRI. Vol. 10950 (International Society for Optics and Photonics, 2019).

12 Jamaspishvili, T. et al. Clinical implications of PTEN loss in prostate cancer. Nat Rev Urol 15, 222–234, doi:10.1038/nrurol.2018.9 (2018).

13 Sinnott, J. A. et al. Prognostic Utility of a New mRNA Expression Signature of Gleason Score. Clin Cancer Res 23, 81–87, doi:10.1158/1078-0432.CCR-16-1245 (2017).

14 Donovan, M. J. et al. Development and validation of a novel automated Gleason grade and molecular profile that define a highly predictive prostate cancer progression algorithm-based test. Prostate Cancer Prostatic Dis 21, 594–603, doi:10.1038/s41391-018-0067-4 (2018).

15 Jeldres, C. et al. Survival after radical prostatectomy and radiotherapy for prostate cancer: a population-based study. Can Urol Assoc J 3, 13–21 (2009).

16 Hervas, A. et al. Outcomes and prognostic factors in intermediate-risk prostate cancer: multi-institutional analysis of the Spanish RECAP database. Clin Transl Oncol 21, 900–909, doi:10.1007/s12094-018-02000-y (2019).

17 Jones, T. A., Radtke, J. P., Hadaschik, B. & Marks, L. S. Optimizing safety and accuracy of prostate biopsy. Curr Opin Urol 26, 472–480, doi:10.1097/MOU.0000000000000310 (2016).

18 Litjens, G. et al. A survey on deep learning in medical image analysis. Med Image Anal 42, 60–88, doi:10.1016/j.media.2017.07.005 (2017).

19 Shen, D., Wu, G. & Suk, H. I. Deep Learning in Medical Image Analysis. Annu Rev Biomed Eng 19, 221–248, doi:10.1146/annurev-bioeng-071516-044442 (2017).

20 Suzuki, K. Overview of deep learning in medical imaging. Radiol Phys Technol 10, 257–273, doi:10.1007/s12194-017-0406-5 (2017).

21 Khosravi, P., Kazemi, E., Imielinski, M., Elemento, O. & Hajirasouliha, I. Deep Convolutional Neural Networks Enable Discrimination of Heterogeneous Digital Pathology Images. EBioMedicine 27, 317–328, doi:10.1016/j.ebiom.2017.12.026 (2018).

22 Khosravi, P. et al. Deep learning enables robust assessment and selection of human blastocysts after in vitro fertilization. npj Digital Medicine 2, 21, doi:https://doi.org/10.1038/s41746-019-0096-y (2019).

23 Wang, X. et al. Searching for prostate cancer by fully automated magnetic resonance imaging classification: deep learning versus non-deep learning. Sci Rep 7, 15415, doi:10.1038/s41598-017-15720-y (2017).

24 Kwon, D. et al. Classification of suspicious lesions on prostate multiparametric MRI using machine learning. J Med Imaging (Bellingham) 5, 034502, doi:10.1117/1.JMI.5.3.034502 (2018).

25 Rubinstein, E. et al. Unsupervised tumor detection in Dynamic PET/CT imaging of the prostate. Med Image Anal 55, 27–40, doi:10.1016/j.media.2019.04.001 (2019).

26 Chiavaras, M. M., Jacobson, J. A., Smith, J. & Dahm, D. L. Pectoralis major tears: anatomy, classification, and diagnosis with ultrasound and MR imaging. Skeletal Radiol 44, 157–164, doi:10.1007/s00256-014-1990-7 (2015).

27 He, K., Zhang, X., Ren, S. & Sun, J. in IEEE conference on computer vision and pattern recognition. 770–778.

28 van der Maaten, L. & Hinton, G. Visualizing Data using t-SNE. J Mach Learn Res 9, 2579–2605 (2008).

29 Esteva, A. et al. Corrigendum: Dermatologist-level classification of skin cancer with deep neural networks. Nature 546, 686, doi:10.1038/nature22985 (2017).

30 Szegedy, C. et al. in Computer Vision and Pattern Recognition (CVPR). 1-9 (IEEE).

31 Zhou, B., Khosla, A., Lapedriza, A., Oliva, A. & Torralba, A. Learning Deep Features for Discriminative Localization. 2016 Ieee Conference on Computer Vision and Pattern Recognition (Cvpr), 2921–2929, doi:10.1109/Cvpr.2016.319 (2016).

32 Woo, S., Kim, S. Y., Cho, J. Y. & Kim, S. H. Preoperative Evaluation of Prostate Cancer Aggressiveness: Using ADC and ADC Ratio in Determining Gleason Score. Am J Roentgenol 207, 114–120, doi:10.2214/Ajr.15.15894 (2016).

33 Yuan, Y. X. et al. Prostate cancer classification with multiparametric MRI transfer learning model. Med Phys 46, 756–765, doi:10.1002/mp.13367 (2019).

34 Cao, R. et al. Joint Prostate Cancer Detection and Gleason Score Prediction in mp-MRI via FocalNet. IEEE Trans Med Imaging, doi:10.1109/TMI.2019.2901928 (2019).

35 Ronneberger, O., Fischer, P. & Brox, T. U-Net: Convolutional Networks for Biomedical Image Segmentation. Medical Image Computing and Computer-Assisted Intervention, Pt Iii 9351, 234–241, doi:10.1007/978-3-319-24574-4_28 (2015).

36 Chen, L. C., Papandreou, G., Kokkinos, I., Murphy, K. & Yuille, A. L. DeepLab: Semantic Image Segmentation with Deep Convolutional Nets, Atrous Convolution, and Fully Connected CRFs. IEEE Trans Pattern Anal Mach Intell 40, 834–848, doi:10.1109/TPAMI.2017.2699184 (2018).

37 Epstein, J. I. et al. The 2014 International Society of Urological Pathology (ISUP) Consensus Conference on Gleason Grading of Prostatic Carcinoma: Definition of Grading Patterns and Proposal for a New Grading System. Am J Surg Pathol 40, 244–252, doi:10.1097/PAS.0000000000000530 (2016).

38 (NCCN), N. C. C. N. Prostate Cancer (Version 4.2018), <https://www.nccn.org/patients/guidelines/prostate/files/assets/common/downloads/files/prostate.pdf> (2018).

39 Szegedy, C., Vanhoucke, V., Ioffe, S., Shlens, J. & Wojna, Z. Rethinking the Inception Architecture for Computer Vision. 2016 Ieee Conference on Computer Vision and Pattern Recognition (Cvpr), 2818–2826, doi:10.1109/Cvpr.2016.308 (2016).

40 Szegedy, C., Ioffe, S., Vanhoucke, V. & Alemi, A. A. Inception-v4, inception-resnet and the impact of residual connections on learning. AAAI, 4278–4284 (2017).

41 Towns, J. et al. XSEDE: Accelerating Scientific Discovery. Comput Sci Eng 16, 62–74, doi:Doi 10.1109/Mcse.2014.80 (2014).

42 Hanley, J. A. & McNeil, B. J. The meaning and use of the area under a receiver operating characteristic (ROC) curve. Radiology 143, 29–36, doi:10.1148/radiology.143.1.7063747 (1982).

43 Zawistowski, M. et al. Corrected ROC analysis for misclassified binary outcomes. Stat Med 36, 2148–2160, doi:10.1002/sim.7260 (2017).

44 Cohen, J. A Coefficient of Agreement for Nominal Scales. Educ Psychol Meas 20, 37–46, doi:Doi 10.1177/001316446002000104 (1960).

